# Impact of FDA’s Enforcement Discretion of Clozapine REMS Program on Clozapine Adverse Event Reporting

**DOI:** 10.1101/2025.04.11.25325669

**Authors:** Nirmal Singh, Zexiang Li, Xin Zhou, Rajiv Radhakrishnan

## Abstract

**Introduction:** In response to the COVID-19 pandemic, the FDA announced enforcement discretion of the clozapine Risk Evaluation and Mitigation Strategy (REMS) program on March 22, 2024. The impact of these changes on rates of Clozapine related adverse-events and reporting pattern has not been examined to-date. Using the FDA Adverse Event Reporting System (FAERS) database, we examined the effect of this regulatory change on the reporting patterns of Clozapine-related adverse events.

**Methods:** Data from the FAERS database from January 2018 to June 2024 was obtained. Interrupted time-series models were employed to evaluate the total number of reported adverse events and event-specific rates for death, agranulocytosis, neutropenia, and myocarditis, with March 22, 2020, as the index date for FDA enforcement discretion.

**Results:** FAERS database reports were predominantly from USA (41.63%), with majority of reports submitted by physicians (37.88%) and pharmacists (37.83%). Total reported counts per month and death count per month did not show significant changes in trend. The reported death proportion and myocarditis count showed a significant downward trend (p = 0.002 and p = 0.005, respectively). No significant trend changes were seen in reporting of agranulocytosis and neutropenia (count per month and proportion).

**Discussion:** The FDA’s enforcement discretion of clozapine REMS program during the COVID-19 pandemic was associated with a significant reduction in reporting of clozapine-induced myocarditis and death, but not of agranulocytosis or neutropenia. Whether these changes reflect true change in rates, preferential attribution of these adverse-events to COVID-19 or change in reporting patterns warrant further verification.

## Introduction

The COVID-19 pandemic presented as a public health emergency between 30 January 2020 – 5 May 2023, and was associated with significant health morbidity and mortality, especially among people with psychiatric disorders^1, 2^ In response to the COVID-19 pandemic, the FDA announced enforcement discretion of the Clozapine Risk Evaluation and Mitigation Strategy (REMS) program on March 22, 2020, allowing for greater flexibility in monitoring requirements^3^. Consequently, the FDA permitted prescribers to use clinical judgment to determine whether the benefits of delaying ANC (absolute neutrophil count) laboratory monitoring outweighed the risks; thus allowing the continuation of Clozapine treatment without updated ANC results ^3^. This regulatory shift was intended to maintain patient safety during the pandemic by minimizing the in-person contact. Further, it allowed the continuation of the clozapine therapy, an atypical antipsychotic approved for treatment-resistant schizophrenia (TRS)^4^.

Approximately 25% of individuals with schizophrenia are affected by treatment-resistant schizophrenia, a condition associated with a worse disease burden and significantly reduced quality of life^5, 6^. TRS remains both prevalent and costly, with healthcare resource utilization costs estimated to be 3 to 11 times higher when compared to the general schizophrenia population^7^. Clozapine remains the drug of choice to treat TRS, with studies showing higher response rate compared to other typical and atypical antipsychotics, including a landmark 1988 study where 30% of TRS patients responded to clozapine versus only 4% with chlorpromazine ^8, 9^.

Despite its merits, clozapine’s use is often limited to treatment-resistant cases due to its safety profile^10^. Clozapine’s FDA label has several potential severe adverse drug reactions warnings including agranulocytosis, neutropenia, and myocarditis. In order to mitigate the severe adverse events, the Clozapine REMS program was established, requiring frequent monitoring of blood count and timely reporting of adverse events ^11–14^. Per the REMS program, following initiation of clozapine, weekly monitoring of white blood cell (WBC) count and absolute neutrophil count (ANC) is required for the first six months, followed by monitoring every 2 weeks from 6 to 12 months, and indefinite monthly monitoring thereafter. This close monitoring requirement adds significant administrative and logistical burden for both the providers and the patients, often dissuading them from prescribing and utilizing clozapine. Further, researchers have suggested that strict monitoring requirements are out of proportion to the risk profile^15^. The incidence of clozapine-induced neutropenia is relatively low, with studies reporting 0.70 cases per 1000 patient-years during months 6 to 12, and decreasing to 0.39 cases per 1000 patient-years after one year^16^. Delayed neutropenia is generally mild, and the risk of fatal neutropenia associated with clozapine after initial exposure is comparable to that of other medications ^17, 18^. Despite these findings, strict monitoring requirements for clozapine and underutilization of its use persists^15^. Recently, in November 2024, a joint meeting of the Drug Safety and Risk Management Advisory Committee (DSRMAC) and the Psychopharmacologic Drugs Advisory Committee (PDAC) convened by the FDA voted against the REMS requirement that prescribers document—and pharmacies verify—adequate ANC values in REMS before dispensing clozapine^19^.

The COVID pandemic and subsequent enforcement discretion to the Clozapine REMS program, provides a naturalistic setting to evaluate the utility and necessity for reporting of ANC and Clozapine-related adverse-events. In this study we examined the impact of the FDA’s enforcement discretion of the Clozapine REMS program on the reporting patterns of Clozapine-related adverse events using the FDA Adverse Event Reporting System (FAERS) database.

## Methods

### Study Sample

The data for this study were sourced from the FAERS database, which is updated quarterly. Data was collated from the first quarter of 2018 (January-March 2018) to the second quarter of 2024 (April-June 2024). The content includes patient demographic information records (DEMO), drug utilization records (DRUG), patient outcome information records (OUTC), and adverse event records (REAC). The search was conducted using “clozapine” as the target product active ingredient. March 22, 2020, was considered as the index date for FDA enforcement discretion.

Data was excluded in a 1-month window on either side of the index date (i.e., Feb 1-April 30, 2020) to account for the time needed for the policy change to take effect. Additionally, COVID-19 cases were excluded to minimize the impact of COVID-19 infections. A sensitivity analysis including data without the 1-month window was also performed. (Results included in Supplementary material)

## Statistical Analysis

Interrupted time-series (ITS) models were employed to analyze the impact of relaxing monitoring guidelines on clozapine-related adverse effects. ITS analysis is a study design for evaluating the effectiveness of population-level health interventions implemented at specific time points. In this study, the outcomes included the total number of reported clozapine-related adverse events per month, as well as specific events such as death, agranulocytosis, myocarditis, and neutropenia associated with clozapine. Additionally, the study also examined the proportions of these adverse events among the total reported clozapine-related adverse events. The intervention was the FDA’s new guidance on relaxation of monitoring on March 22, 2020. The time scale of the analysis is measured in months since the start of the study. The ITS models are shown as the following linear models,

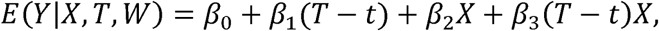

where T is the month elapsed since the start of the study, X is a dichotomous variable for the intervention, indicating the pre-intervention period (coded 0) or the post-intervention period (coded 1), and t is the month starting the intervention. Furthermore, after fitting the model, Cook’s distance, a measure that quantitatively identifies influential data points on the estimates, was used to assess the presence of potential outliers. Data points with Cook’s distance exceeding the conventional threshold 1 were considered influential outliers and excluded. All statistical tests were carried out according to the refitted ITS model by F tests.

## Results

52526 (65.85%) initial reports and 27244 follow-up reports in the FAERS dataset were included in the analysis. The study population had a mean age of 47.98±17.51 years. The gender distribution was 34.32% females and 53.64% males, with 0.05% reporting an unknown sex and 11.98% of data missing. The clozapine adverse effects reports can be submitted by various individuals. In the dataset, 37.88% of reports were by physicians, 37.83% by pharmacists, 13.32% by consumers, and 9.5% by other healthcare professionals. Within the data, 41.63% were reported from the United States, 20.06% from Great Britain, 12.11% from Canada, 5.69% from Australia, and the remaining 20.51% from other countries. The majority of the reports (92.12%) were expedited, while 7.40% were periodic reports.

In our analysis, no data-points had a Cook’s distance exceeding 1, suggesting that no potential outliers impacted the results. The ITS model for the total count (i.e. total number of reports) showed that after the index date (i.e., after FDA enforcement discretion of Clozapine REMS program) there was a trend towards increase in the reported total count, although this change was not significant (Figure 1, p=0.72).

**Figure 1:**
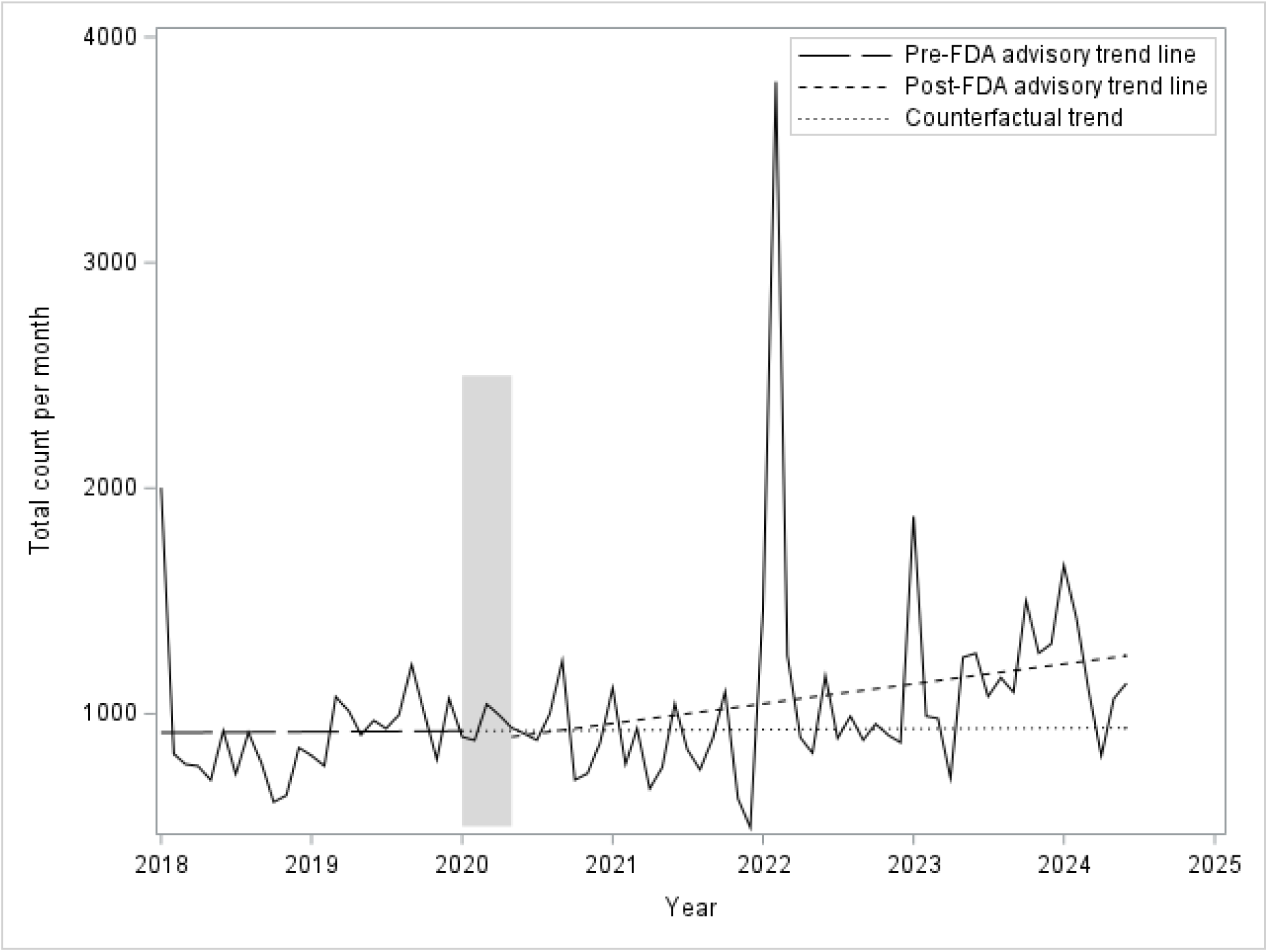
Results of interrupted time series for total count per month β**_1_:** 0.281, β**_2_:**-40.140, β**_3_:** 7.041, **F**: 0.33, **p-value:** 0.718 β**_1_:** Slope of the trend before intervention β**_2_:** Change in outcome level at the time of intervention β**_3_:** Change in slope after the intervention Shaded bar indicates the excluded time window around the index date (i.e. Feb 1-April 30, 2020).

In the ITS model for death count, there was no significant change in reported deaths per month (Figure 2, p=0.14). However, the model for reported death proportion (proportion of total count) revealed a significant downward trend after the index date (Figure 3, p = 0.002) probably due to the increasing number of total counts as noted above.

**Figure 2:**
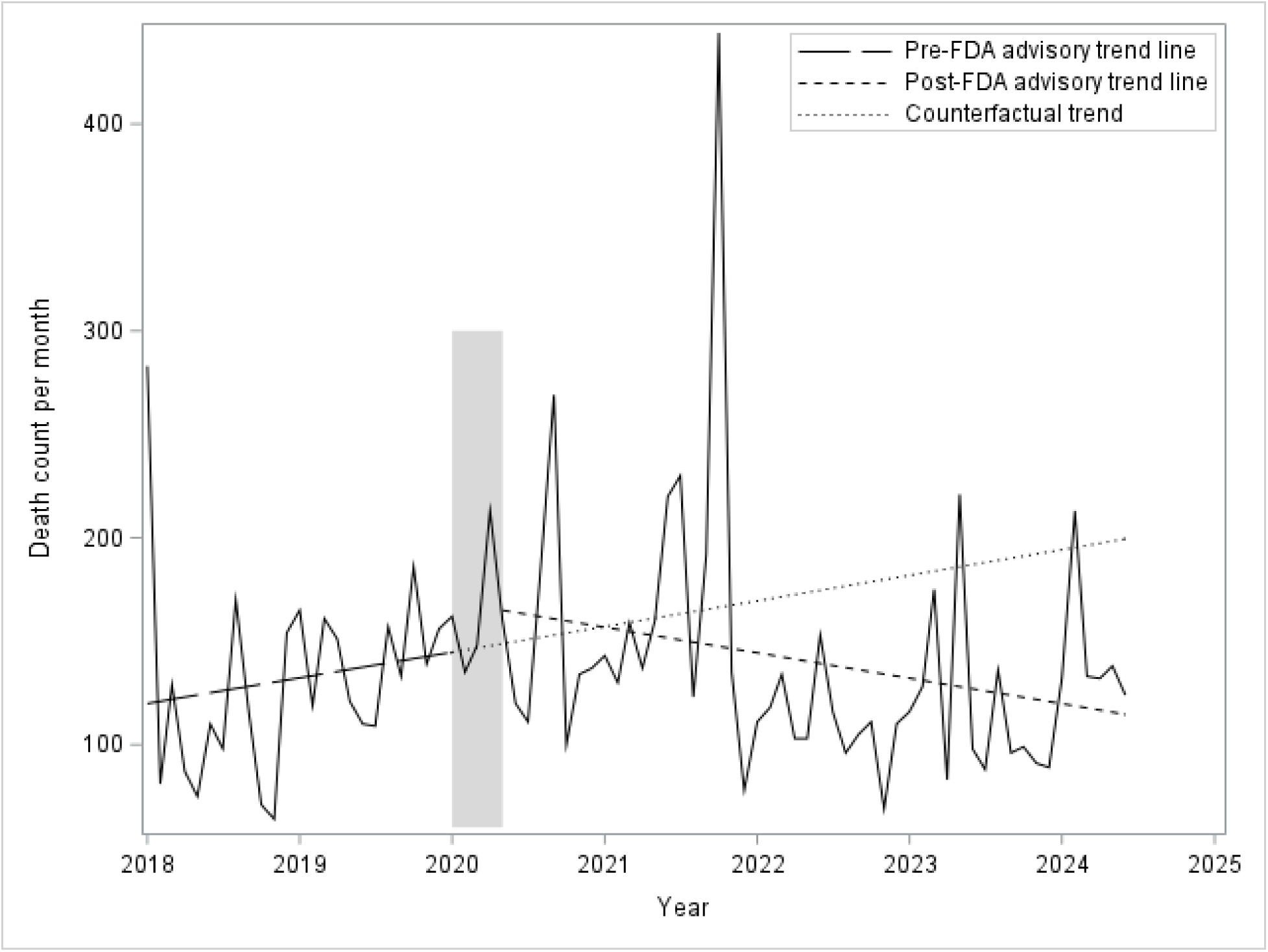
Results of interrupted time series for death count per month β**_1_:** 1.032, β**_2_:** 20.254, β**_3_:** 2.058, **F**: 1.06, **p-value:** 0.136 β**_1_:** Slope of the trend before intervention β**_2_:** Change in outcome level at the time of intervention β**_3_:** Change in slope after the intervention Shaded bar indicates the excluded time window around the index date (i.e. Feb 1-April 30, 2020).

**Figure 3:**
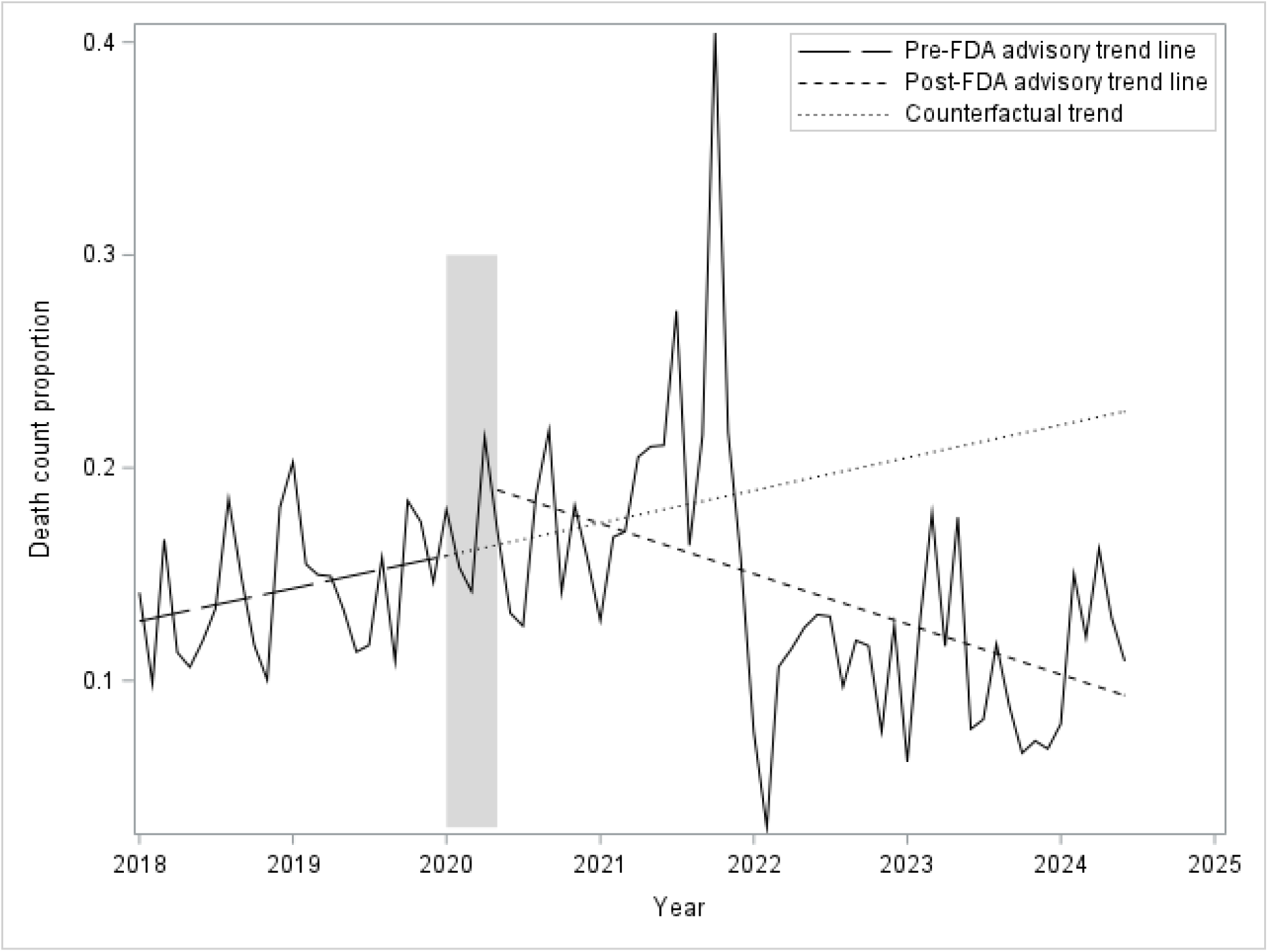
Results of interrupted time series death count proportion β**_1_:** 0.001, β**_2_:** 0.032, β**_3_:**-0.003**, **F**: 6.56, **p-value:** 0.002* β**_1_:** Slope of the trend before intervention β**_2_:** Change in outcome level at the time of intervention β**_3_:** Change in slope after the intervention Shaded bar indicates the excluded time window around the index date (i.e. Feb 1-April 30, 2020).

For reported agranulocytosis cases per month, the total numbers were small, and the ITS model did not show a significant change in trend after the index date (Figure 4, p = 0.34). Similarly, the models did not show a significant change in reported agranulocytosis proportion (proportion of the total count) after the index date (Figure 5, p = 0.07). With regard to myocarditis, there was a significant decrease in the number of myocarditis cases after the index date (Figure 6, p = 0.0078). However, the reported myocarditis proportion did not show a significant change in trend (Figure 7, p = 0.10), although the proportion was lower after the index date. For neutropenia, there was no significant change in both reported neutropenia cases (Figure 8, p = 0.86) and reported neutropenia proportion (Figure 9, p = 0.64) in our analysis.

**Figure 4:**
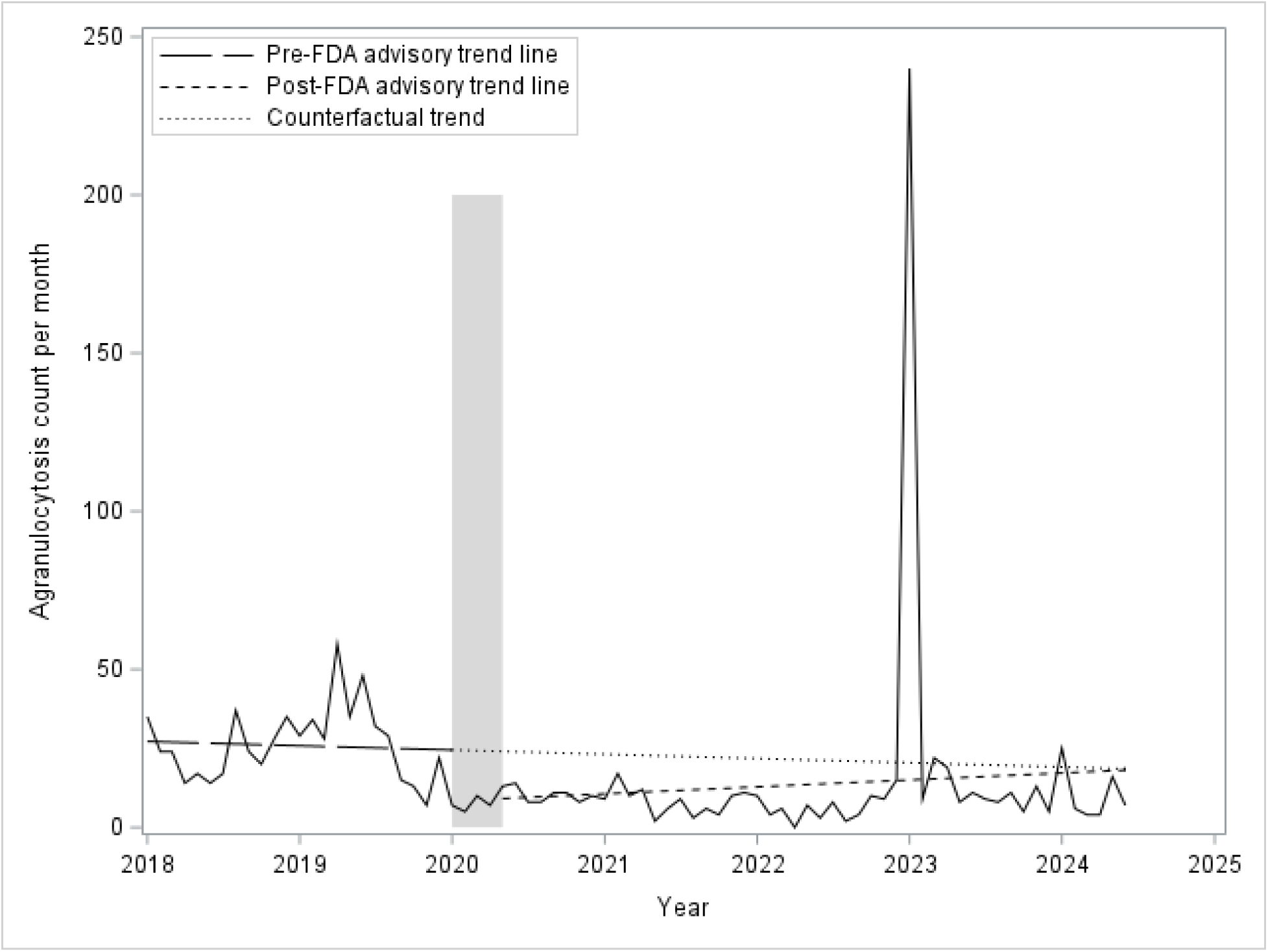
Results of interrupted time series agranulocytosis count per month β**_1_:** 0.112, β**_2_:**-15.466, β**_3_:** 0.295, **F**: 1.11, **p-value:** 0.336 β**_1_:** Slope of the trend before intervention β**_2_:** Change in outcome level at the time of intervention β**_3_:** Change in slope after the intervention Shaded bar indicates the excluded time window around the index date (i.e. Feb 1-April 30, 2020).

**Figure 5:**
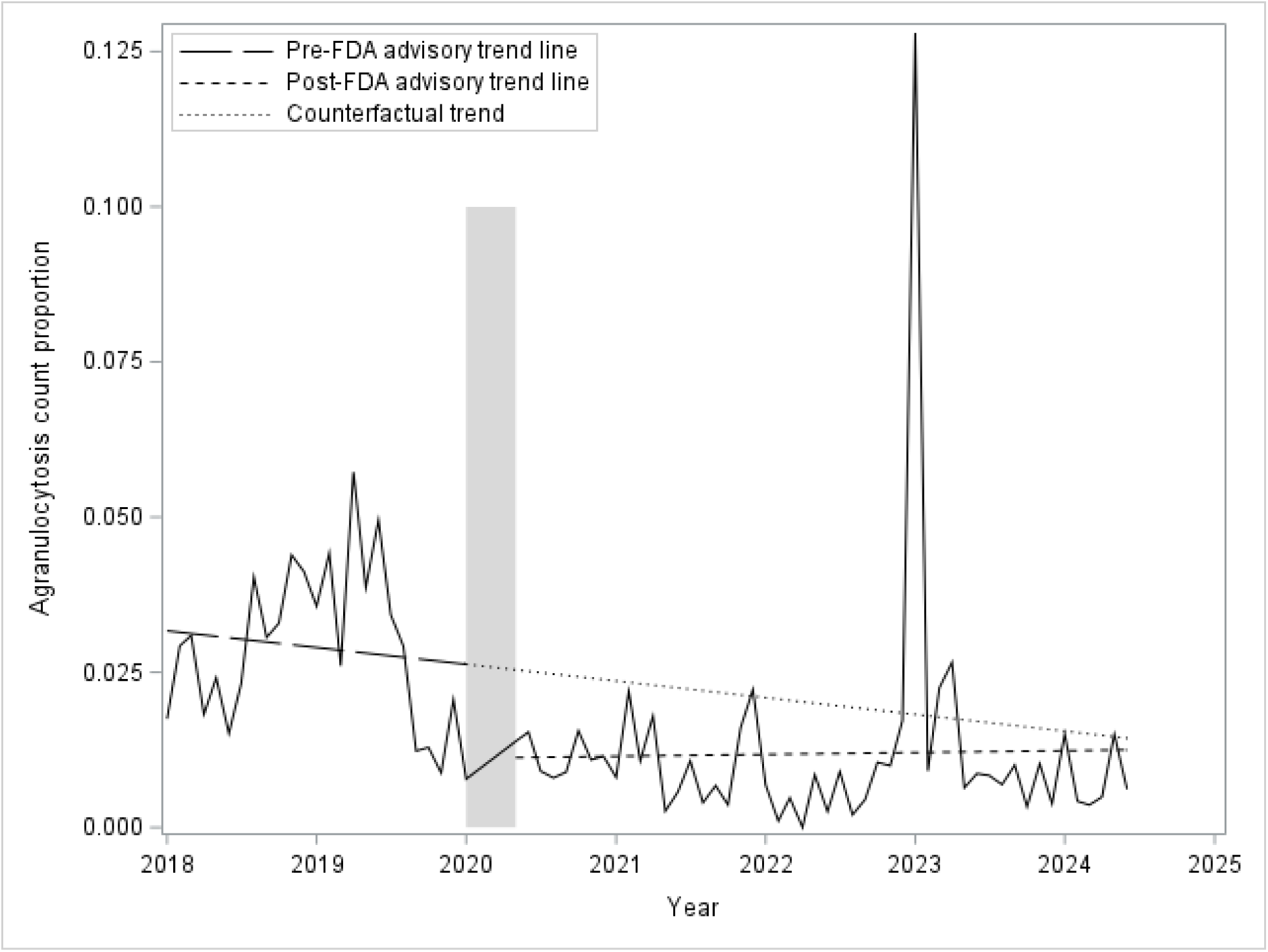
Results of interrupted time series agranulocytosis count proportion β**_1_:** 0.000, β**_2_:**-0.015, β**_3_:** 0.002, **F**: 2.75, **p-value:** 0.070 β**_1_:** Slope of the trend before intervention β**_2_:** Change in outcome level at the time of intervention β**_3_:** Change in slope after the intervention Shaded bar indicates the excluded time window around the index date (i.e. Feb 1-April 30, 2020).

**Figure 6:**
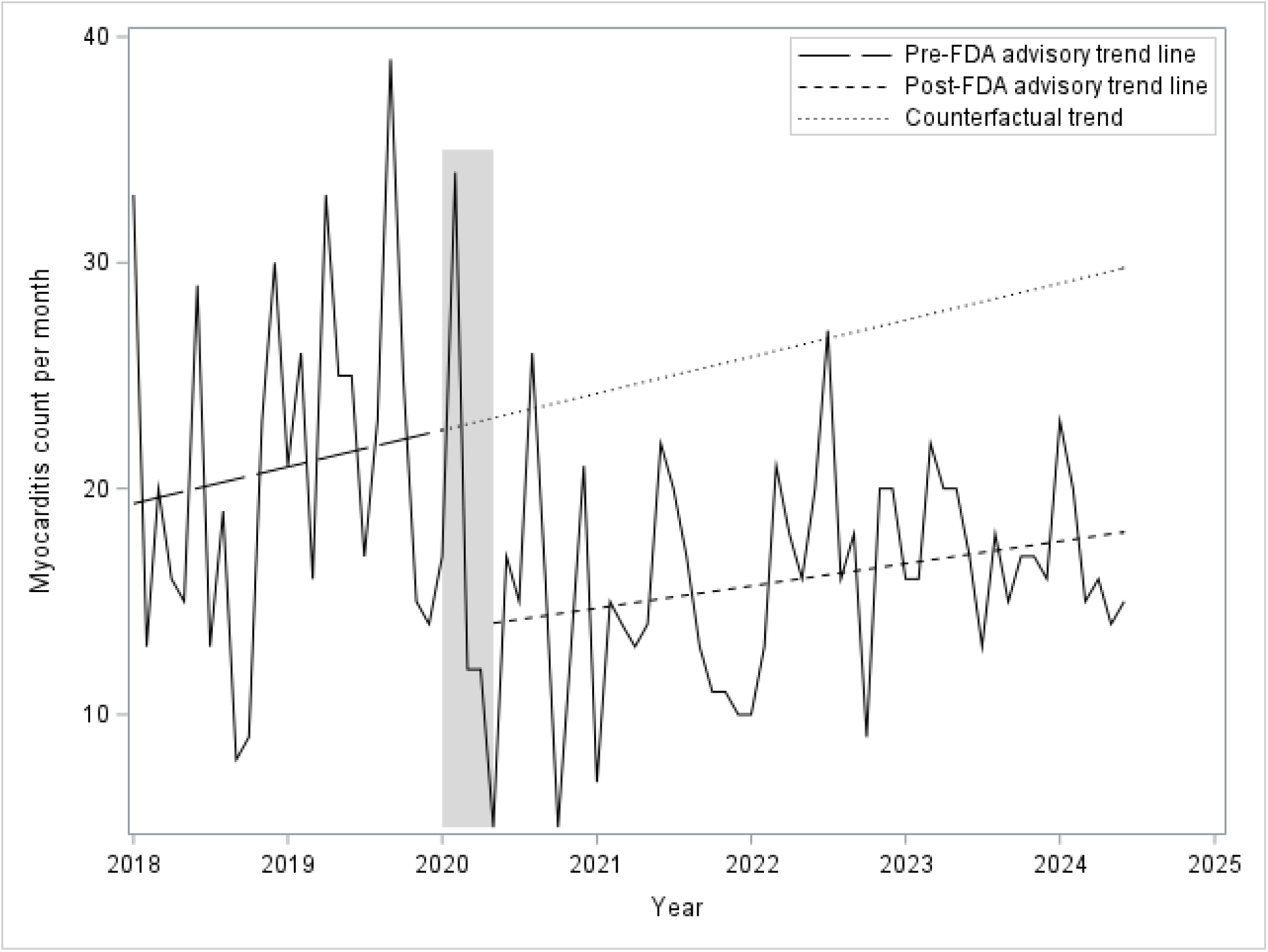
Results of interrupted time series myocarditis count per month β1: 0.135, β2:-8.976**, β3:-0.053, F: 5.20, p-value: 0.007** β**_1_:** Slope of the trend before intervention β**_2_:** Change in outcome level at the time of intervention β**_3_:** Change in slope after the intervention Shaded bar indicates the excluded time window around the index date (i.e. Feb 1-April 30, 2020).

**Figure 7:**
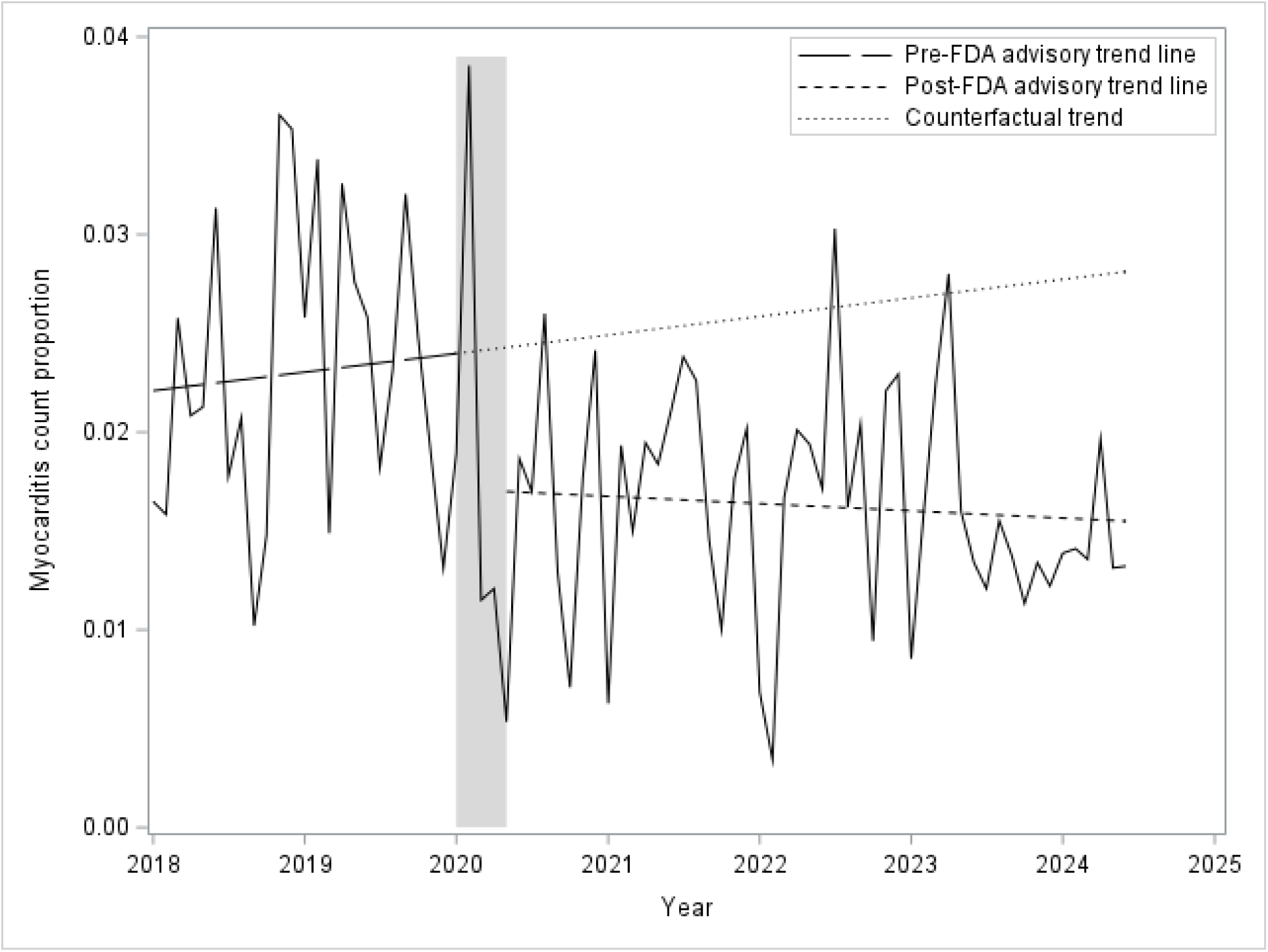
Results of interrupted time series myocarditis count proportion β1: 0.000, β2:-0.007, β3: 0.0001, F: 2.34, p-value: 0.103 β**_1_:** Slope of the trend before intervention β**_2_:** Change in outcome level at the time of intervention β**_3_:** Change in slope after the intervention Shaded bar indicates the excluded time window around the index date (i.e. Feb 1-April 30, 2020).

**Figure 8:**
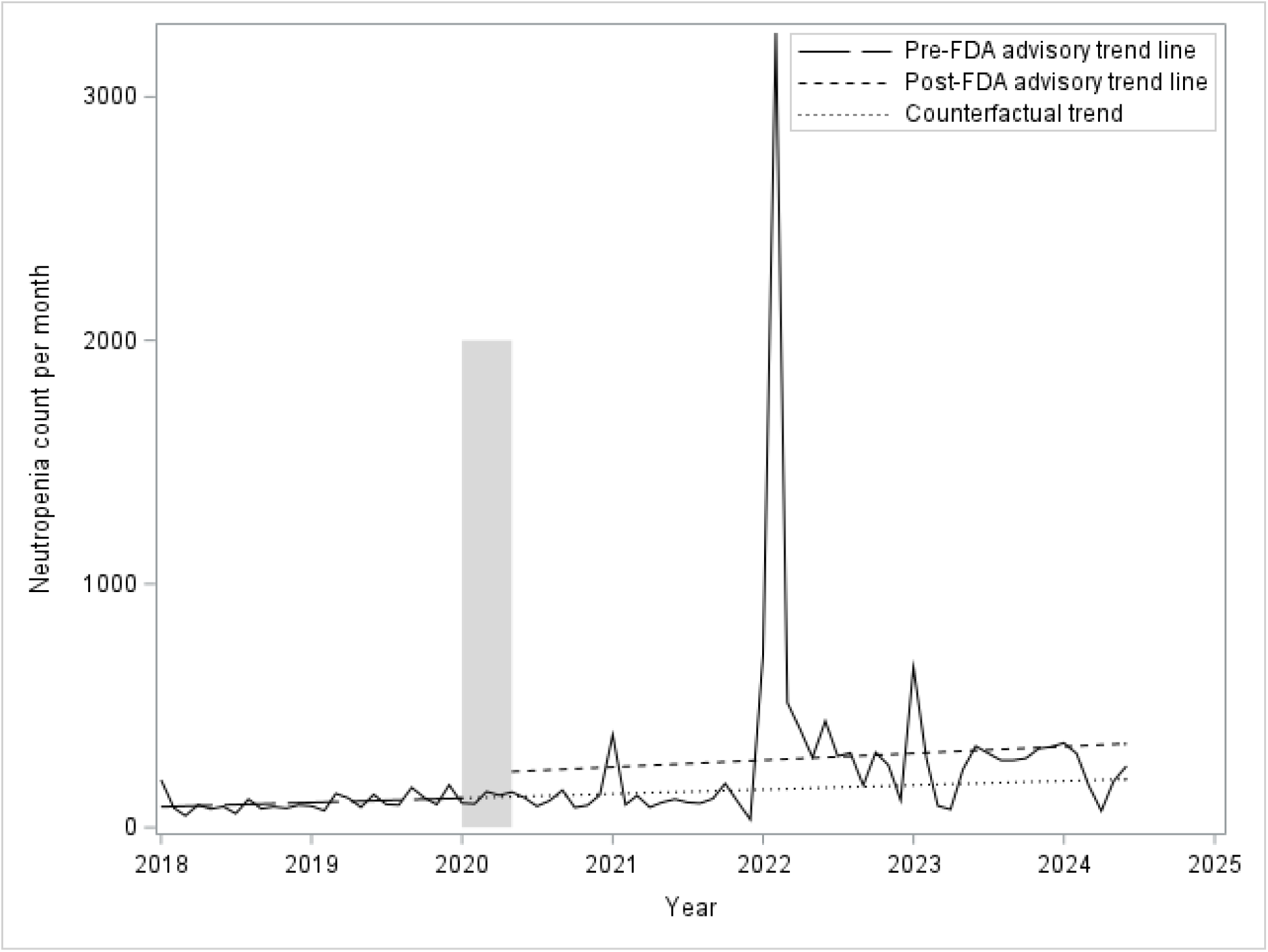
Results of interrupted time series neutropenia count per month β1: 1.476, β2: 101.332, β3: 0.873, F: 0.16, p-value: 0.856 β**_1_:** Slope of the trend before intervention β**_2_:** Change in outcome level at the time of intervention β**_3_:** Change in slope after the intervention Shaded bar indicates the excluded time window around the index date (i.e. Feb 1-April 30, 2020).

**Figure 9:**
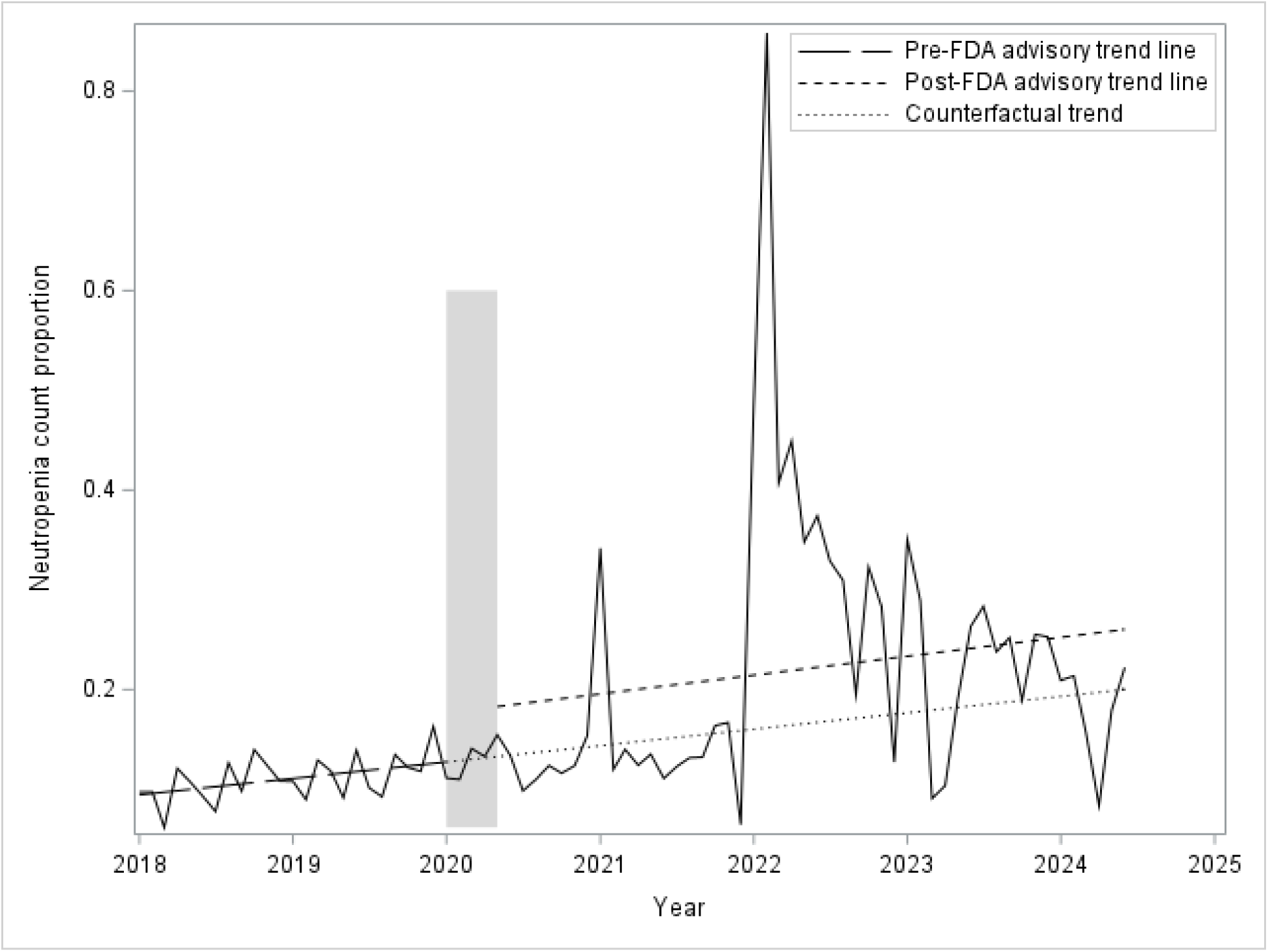
Results of interrupted time series neutropenia count proportion β1: 0.007, β2: 0.089, β3: 0.001, F: 0.45, p-value: 0.681 β**_1_:** Slope of the trend before intervention β**_2_:** Change in outcome level at the time of intervention β**_3_:** Change in slope after the intervention Shaded bar indicates the excluded time window around the index date (i.e. Feb 1-April 30, 2020).

## Discussion

This is the first study, to our knowledge, to examine the impact of FDA’s temporary enforcement discretion of Clozapine REMS program on the reporting of clozapine-related adverse events. The COVID-19 pandemic provided a unique opportunity to evaluate the impact of relaxation of monitoring requirements on the reporting of clozapine-related adverse events. Our analysis did not reveal a significant change in reported counts of death, neutropenia, or agranulocytosis.

However, we found that the reporting of death proportion was significantly lower following FDA’s enforcement discretion, probably explained by an increase in total number of events reported. There was no difference in the proportion of neutropenia or agranulocytosis. The number of myocarditis cases reported was significantly lower following FDA’s enforcement discretion although there was no difference in the proportion of myocarditis. These changes might reflect a true change in rates, preferential attribution of these adverse-events to COVID-19, or a change in reporting patterns. The relationship between COVID-19 and myocarditis has been well-documented, and COVID-19 vaccination has been associated with higher rates of myocarditis, although rates of myocarditis were also increased in the COVID-19 infections^20^.

Our finding contributes to the ongoing discussion surrounding monitoring of clozapine-related adverse events. There is some literature supporting less stringent monitoring. A large London NHS trust reduced monitoring intervals from four-weekly to 12-weekly during the COVID-19 pandemic, reporting no increase in severe neutropenia during this period^21^. Along the similar lines, global study of 102 countries found that only 42 countries enforce mandatory monitoring^12^. Some countries such as Bulgaria, Mexico, and Colombia limit their hematological monitoring to the first month of treatment. In Iceland, monitoring occurs approximately every four months, while in the Netherlands, patients with capacity can request to cease monitoring entirely^12^. These findings highlight that international practices vary significantly, with some less stringent approaches proving safe and effective. In line with these findings, a consensus statement by Siskind et al. recommended less frequent monitoring during the pandemic, provided patients were carefully selected and closely monitored for signs of infection or clozapine toxicity^4^.

Taking a more longitudinal view of the clozapine related adverse events show that rates of adverse events vary as a function of time since the start of the treatment. Clozapine-induced myocarditis has a global incidence of approximately 0.1%. Notably higher rate of ∼10 times in Australia were reported which is postulated due to underreporting and irregular monitoring in the rest of the world ^22–24^.Clozapine is associated with a higher risk of myocarditis compared to chlorpromazine and other antipsychotic medications^23, 25^. As the peak risk of clozapine-induced myocarditis (CIM) is around 3 months, during this time close monitoring and early detection through symptoms and signs reduce the risk of mortality ^23, 26^.

Clozapine associated agranulocytosis (CIA) and neutropenia (CIN) are hematological side-effects requiring close monitoring. CIA occurs in 0.7-0.9% of patients and usually peaks in the third month of treatment^17, 27^. Some evidence suggests that it rarely occurs after the first year.^28^

After the first year, the risk of CIA is comparable to other antipsychotics with some studies even reporting lower risk with clozapine^18^. CIN occurs in approximately 3-3.8% of patients and can occur at any time^17^.

Fixed regular monitoring and reporting is thought to add significant burden and create barriers to prescription of clozapine^29^. REMS requirements have been reported to often lead to underutilization and treatment interruptions. In order to address these concerns, the FDA convened a panel which voted 14-1 to eliminate the REMS requirements^19^. On February 24, 2025, FDA discontinued the requirement for prescribers, pharmacies, and patients to participate in the clozapine REMS or to report ANC results before pharmacies dispense clozapine ^30^. It is important to note that this does not change physician practice regarding ANC monitoring. FDA still recommends that prescribers monitor patients’ ANC according to the monitoring frequencies described in the prescribing information.

Although the requirement of ANC monitoring adds significant economic and administrative cost for the patients and providers, there are some benefits to this practice. Monitoring promotes regular engagement with the healthcare system which in turn leads to enhanced therapeutic alliance, medication adherence and regular health checkups. The regular health check-ups are also helpful in early detection of more severe adverse events such as myocarditis and bowel obstruction.

It is important to consider the limitations of the FAERS dataset in interpreting these findings. The FAERS dataset is not designed to monitor temporal changes in reporting patterns and is susceptible to duplicate entries. While there were no outliers identified by Cook’s distance, there were periods with very high and very low adverse events reported. Thus, it is important to consider the possibilities of underreporting and irregular data reporting during the pandemic, as well as the potential for COVID-19 itself to have influenced adverse event proportions.

In conclusion, FDA’s temporary enforcement discretion of Clozapine REMS program did not result in an increase in reported adverse events among clozapine-treated patients. Nevertheless, individualized risk assessment and vigilance in adverse event monitoring remain critical. These findings contribute to the ongoing discussion about regulatory requirements and the balance between patient safety and practical considerations during public health emergencies. Future research should focus on assessing the long-term impacts of regulatory changes on patient outcomes and identifying patient populations that may safely benefit from adjusted monitoring protocols.

## Data Availability

Data is obtained from the publicly available FAER's dataset.

---

**Table 1:**
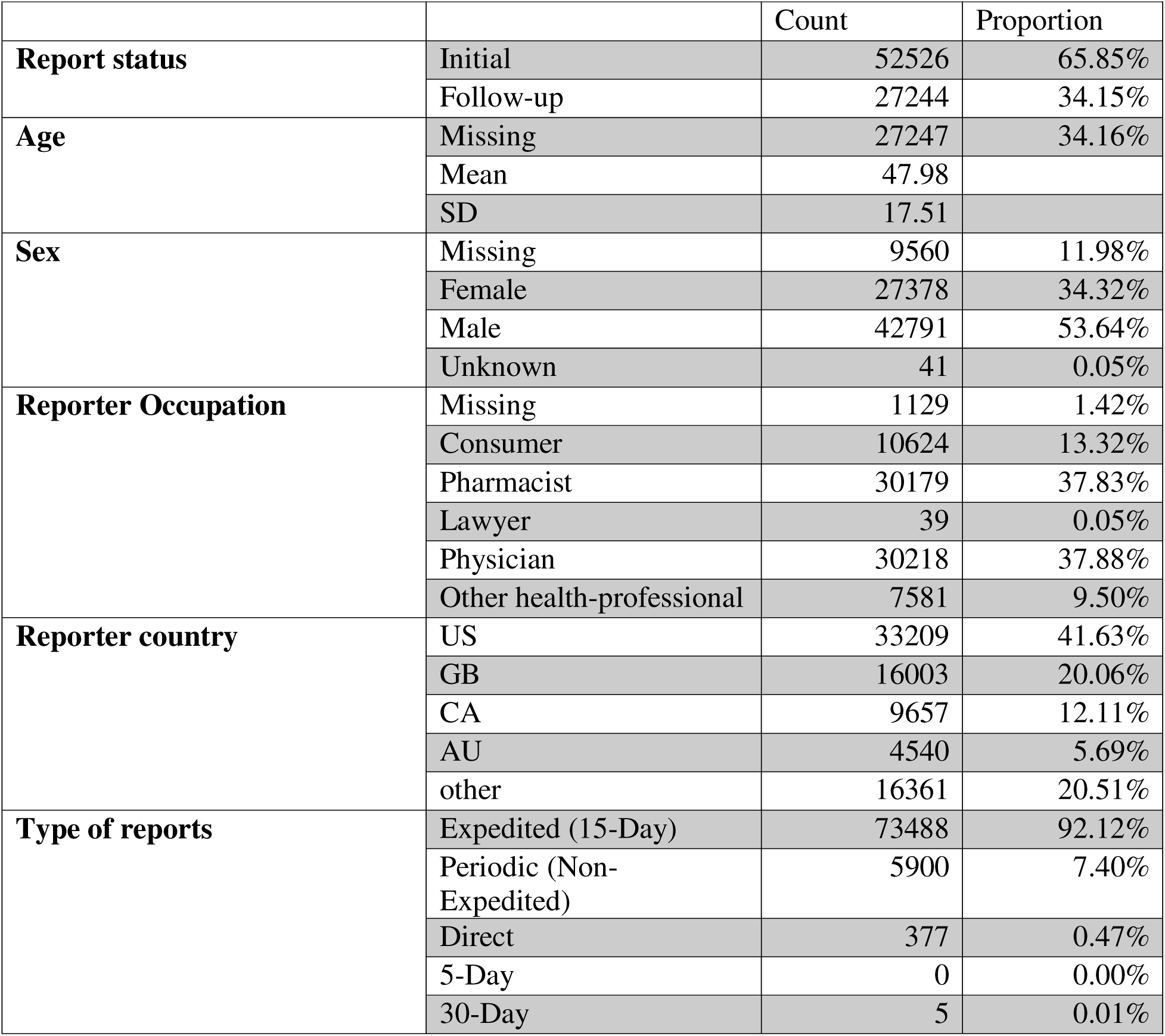
Demographics.

**Table 2:** Change in trend of reported events following FDA Enforcement discretion of REMS program.

| Outcomes | Slope of the trend before intervention ($\beta_1$) | Change in the outcome at the time of intervention ($\beta_2$) | Change in slope after the intervention ($\beta_3$) | Overall test intervention results | |
| --- | --- | --- | --- | --- | --- |
|  |  |  |  | F | p |
| **Total Count** | 0.281 | -40.140 | 7.041 | 0.33 | 0.718 |
| **Death Count** | 1.032 | 20.254 | 2.058 | 2.06 | 0.136 |
| **Death Proportion** | 0.001 | 0.032 | -0.003** | 6.56 | 0.002** |
| **Agranulocytosis Count** | 0.112 | -15.466 | 0.295 | 1.11 | 0.336 |
| **Agranulocytosis Proportion** | -0.000 | -0.015 | 0.0002 | 2.75 | 0.070 |
| **Myocarditis Count** | 0.135 | -8.976** | -0.053 | 5.20 | 0.007** |
| **Myocarditis Proportion** | 0.000 | -0.007** | -0.0001 | 2.34 | 0.103 |
| **Neutropenia Count** | 1.476 | 101.332 | 0.873 | 0.16 | 0.856 |
| **Neutropenia Proportion** | 0.001 | 0.049 | 0.001 | 0.45 | 0.641 |

\*\* indicates statistical significance (p-value < 0.05)

## References

1. Christian NJ, Zhou X, Radhakrishnan R. Effects of Buprenorphine, Methadone, and Substance Use on COVID-19 Morbidity and Mortality. J Addict Med 2025 Mar-Apr 01;19(2):223-226.

2. Ramakrishnan D, Sureshanand S, Pittman B, Radhakrishnan R. Impact of Cannabis Use, Substance Use Disorders, and Psychiatric Diagnoses on COVID-19 Outcomes: A Retrospective Cohort Study. J Clin Psychiatry 2022 Aug 29;83(5).

3. FDA. FDA provides update on patient access to certain REMS drugs during COVID-19 public health emergency [Internet]. FDA. 2020 Mar 22 [cited 2020 Apr 8]. Available from: https://www.fda.gov/news-events/press-announcements/coronavirus-covid-19-update-fda-provides-update-patient-access-certain-rems-drugs-during-covid-19. 2020.

4. Siskind D, Honer W, Clark S, et al. Consensus statement on the use of clozapine during the COVID-19 pandemic. JOURNAL OF PSYCHIATRY & NEUROSCIENCE 2020 2020-05-01;45:222-223.

5. Siskind D, Orr S, Sinha S, et al. Rates of treatment-resistant schizophrenia from first-episode cohorts: systematic review and meta-analysis. BRITISH JOURNAL OF PSYCHIATRY 2022 2021-05-11;220:115-120.

6. Radhakrishnan R, Ganesh S, Meltzer HY, et al. Schizophrenia. In: Ebert MH, Leckman JF, Petrakis IL, eds. Current Diagnosis &Treatment: Psychiatry, 3e. New York, NY: McGraw-Hill Education; 2019.

7. Kennedy J, Altar C, Taylor D, Degtiar I, Hornberger J. The social and economic burden of treatment-resistant schizophrenia: a systematic literature review. INTERNATIONAL CLINICAL PSYCHOPHARMACOLOGY 2014 2014-03-01;29:63-76.

8. Kane J, Honigfeld G, Singer J, Meltzer H. CLOZAPINE FOR THE TREATMENT-RESISTANT SCHIZOPHRENIC - A DOUBLE-BLIND COMPARISON WITH CHLORPROMAZINE. Archives of General Psychiatry 1988 Sep;45(9):789–796.

9. Stroup T, Gerhard T, Crystal S, Huang C, Olfson M. Comparative Effectiveness of Clozapine and Standard Antipsychotic Treatment in Adults With Schizophrenia. AMERICAN JOURNAL OF PSYCHIATRY 2016 2016-02-01;173:166-173.

10. Miller D. Review and management of clozapine side effects. JOURNAL OF CLINICAL PSYCHIATRY 2000 2000-01-01;61:14-17.

11. Clozaril Package Insert. [cited; Available from: /https://www.accessdata.fda.gov/drugsatfda_docs/label/2017/019758s084lbl.pdf

12. Oloyede E, Blackman G, Whiskey E, et al. Clozapine haematological monitoring for neutropenia: a global perspective. EPIDEMIOLOGY AND PSYCHIATRIC SCIENCES 2022 2022-11-25;31.

13. Oloyede E, Taylor D, MacCabe J. International Variation in Clozapine Hematologic Monitoring-A Call for Action. JAMA PSYCHIATRY 2023 2023-04-05;80:535-536.

14. FDA. Guidance for industry: format and content of proposed Risk Evaluation and Mitigation Strategies (REMS), REMS assessments, and proposed REMS modifications: draft guidance [Internet]. October 2017 2017 [cited; Available from: http://www.fda.gov/downloads/Drugs/GuidanceCompliance.RegulatoryInformation/Guidances/UCM184128.pdf

15. Baig A, Bazargan-Hejazi S, Ebrahim G, Rodriguez-Lara J. Clozapine prescribing barriers in the management of treatment-resistant schizophrenia A systematic review. MEDICINE 2021 2021-11-12;100.

16. Schulte P. Risk of clozapine-associated agranulocytosis and mandatory white blood cell monitoring. ANNALS OF PHARMACOTHERAPY 2006 2006-04-01;40:683-688.

17. Myles N, Myles H, Xia S, et al. Meta-analysis examining the epidemiology of clozapine-associated neutropenia. ACTA PSYCHIATRICA SCANDINAVICA 2018 2018-08-01;138:101-109.

18. Myles N, Myles H, Xia S, et al. A meta-analysis of controlled studies comparing the association between clozapine and other antipsychotic medications and the development of neutropenia. AUSTRALIAN AND NEW ZEALAND JOURNAL OF PSYCHIATRY 2019 2019-05-01;53:403-412.

19. Richmond LM. FDA Panel Votes Overwhelmingly to Abolish Clozapine REMS. American Psychiatric Publishing, Inc.; 2025.

20. Heidecker B, Dagan N, Balicer R, et al. Myocarditis following COVID-19 vaccine: incidence, presentation, diagnosis, pathophysiology, therapy, and outcomes put into perspective. A clinical consensus document supported by the Heart Failure Association of the European Society of Cardiology (ESC) and the ESC Working Group on Myocardial and Pericardial Diseases. European journal of heart failure 2022;24(11):2000–2018.

21. Oloyede E, Dzahini O, Abolou Z, et al. Clinical impact of reducing the frequency of clozapine monitoring: controlled mirror-image cohort study. BRITISH JOURNAL OF PSYCHIATRY 2023 2023-04-24;223(2):382-388.

22. Curto M, Girardi N, Lionetto L, Ciavarella G, Ferracuti S, Baldessarini R. Systematic Review of Clozapine Cardiotoxicity. CURRENT PSYCHIATRY REPORTS 2016 2016-07-01;18(7).

23. Bellissima B, Tingle M, Cicovic A, Alawami M, Kenedi C. A systematic review of clozapine-induced myocarditis. INTERNATIONAL JOURNAL OF CARDIOLOGY 2018 2018-05-15;259:122-129.

24. Reinders J, Parsonage W, Lange D, Potter J, Plever S. Clozapine-related myocarditis and cardiomyopathy in an Australian metropolitan psychiatric service. AUSTRALIAN AND NEW ZEALAND JOURNAL OF PSYCHIATRY 2004 2004-11-01;38(11-12):915-922.

25. Coulter D, Bate A, Meyboom R, Lindquist M, Edwards I. Antipsychotic drugs and heart muscle disorder in international pharmacovigilance: data mining study. BMJ-BRITISH MEDICAL JOURNAL 2001 2001-05-19;322(7296):1207-1209.

26. Merrill D, Dec G, Goff D. Adverse cardiac effects associated with clozapine. JOURNAL OF CLINICAL PSYCHOPHARMACOLOGY 2005 2005-02-01;25(1):32-41.

27. Zimbron J, Khandaker G, Toschi C, Jones P, Fernandez-Egea E. A systematic review and meta-analysis of randomised controlled trials of treatments for clozapine-induced obesity and metabolic syndrome. EUROPEAN NEUROPSYCHOPHARMACOLOGY 2016 2016-09-01;26(9):1353-1365.

28. Roge R, Moller B, Andersen C, Correll C, Nielsen J. Immunomodulatory effects of clozapine and their clinical implications: What have we learned so far? SCHIZOPHRENIA RESEARCH 2012 2012-09-01;140(1-3):204-213.

29. Wicinski M, Weclewicz M. Clozapine-induced agranulocytosis/granulocytopenia: mechanisms and monitoring. CURRENT OPINION IN HEMATOLOGY 2018 2018-01-01;25(1):22-28.

30. FDA. Information on Clozapine. 2025.

