## Supplemental for "Impact of FDA’s Enforcement Discretion of Clozapine REMS Program on Clozapine Adverse Event Reporting"

**Supplementary tables and figures:**

**Content:**

**Supplementary Table 1:**  Change in trend of reported events following FDA Enforcement discretion of REMS program – analysis with 2-month window around index date

**Supplementary Figure 1:** Results of interrupted time series for total count per month (analysis including 2-month window around index date)

**Supplementary Figure 2:** Results of interrupted time series for death count per month (analysis including 2-month window around index date)

**Supplementary Figure 3:** Results of interrupted time series death count proportion (analysis including 2-month window around index date)

**Supplementary Figure 4:** Results of interrupted time series agranulocytosis count per month (analysis including 2-month window around index date)

**Supplementary Figure 5:** Results of interrupted time series agranulocytosis count proportion (analysis including 2-month window around index date)

**Supplementary Figure 6:** Results of interrupted time series myocarditis count per month (analysis including 2-month window around index date)

**Supplementary Figure 7:** Results of interrupted time series myocarditis count proportion (analysis including 2-month window around index date)

**Supplementary Figure 8:** Results of interrupted time series neutropenia count per month (analysis including 2-month window around index date)

**Supplementary Figure 9:** Results of interrupted time series neutropenia count proportion (analysis including 2-month window around index date)

**Abbreviation:**

**β_1_:** Slope of the trend before intervention

**β_2_:** Change in outcome level at the time of intervention

**β_3_:** Change in slope after the intervention

**Supplementary Table 1:** Change in trend of reported events following FDA Enforcement discretion of REMS program – analysis with 2-month window around index date

|  | Slope of the trend before intervention (β_1_) | Change in outcome level at the time of intervention (β_2_) | Change in slope after the intervention (β_3_) | Overall test intervention results | |
| --- | --- | --- | --- | --- | --- |
|  |  |  |  | **F** | **p** |
| **Total Count** | 0.282 | -104.319 | 7.722 | 1.81 | 0.171 |
| **Death Count** | 1.032 | 23.416 | -2.104 | 2.30 | 0.108 |
| **Death Proportion** | 0.002 | 0.036 | -0.003**** | 7.44 | 0.001** |
| **Agranulocytosis Count** | 0.112 | -15.135 | 0.291 | 1.02 | 0.365 |
| **Agranulocytosis Proportion** | -0.0002 | -0.015 | 0.0002 | 2.69 | 0.075 |
| **Myocarditis Count** | 0.135 | -8.692 | -0.058 | 4.71 | 0.012 |
| **Myocarditis Proportion** | 0.00007 | -0.007**** | -0.0001 | 2.20 | 0.118 |
| **Neutropenia Count** | 1.476 | 19.400 | 1.801 | 0.14 | 0.870 |
| **Neutropenia Proportion** | 0.001 | 0.0354 | 0.0003 | 0.39 | 0.681 |

** indicates statistical significance (p-value < 0.05)

**Supplementary Figure 1:** Results of interrupted time series for total count per month ( analysis including 2-month window around index date)

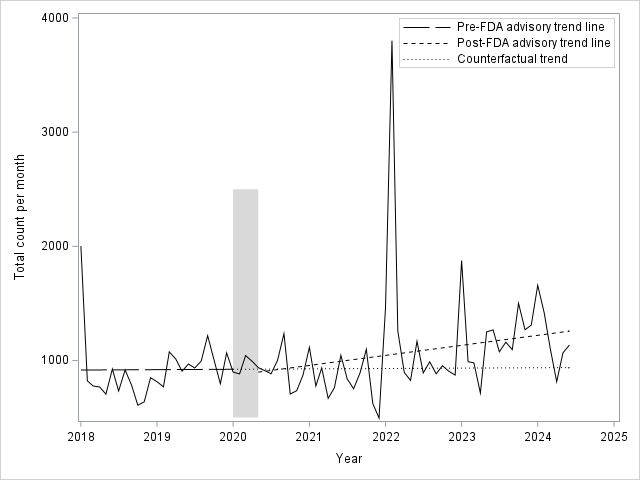

**β_1_:** 0.282, **β_2_:** -104.319, **β_3_:** 7.722, **F**: 1.81, **p-value:** 0.171

**Supplementary Figure 2:** Results of interrupted time series for death count per month (analysis including 2-month window around index date)

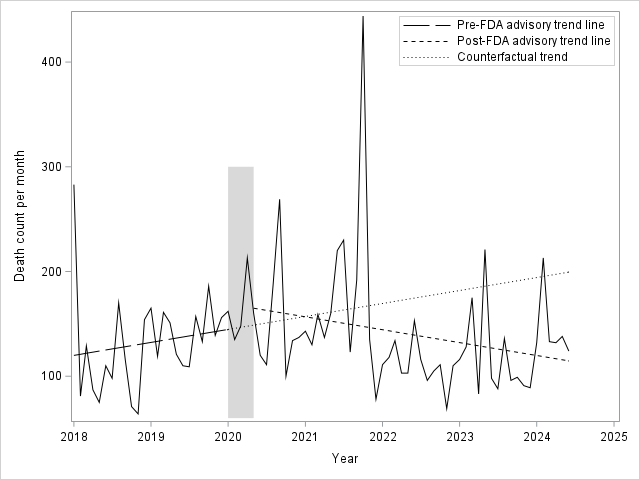

**β_1_:** 1.032, **β_2_:** 23.416, **β_3_:** -2.104, **F**: 2.30, **p-value:** 0.108

**Supplementary Figure 3:** Results of interrupted time series death count proportion (analysis including 2-month window around index date)

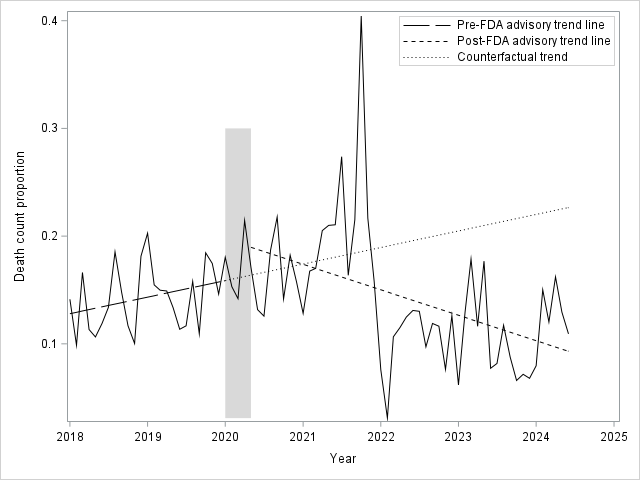

**β_1_**0.002, **β_2_:** 0.036, **β_3_:** -0.003****, **F**: 7.44, **p-value:** 0.001**

**Supplementary Figure 4:** Results of interrupted time series agranulocytosis count per month (analysis including 2-month window around index date)

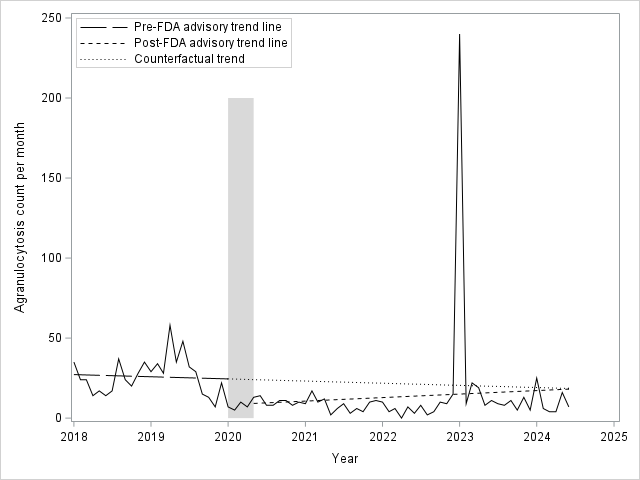

**β_1_:** 0.112, **β_2_:** -15.135, **β_3_:** 0.291, **F**: 1.02, **p-value:** 0.365

**Supplementary Figure 5:** Results of interrupted time series agranulocytosis count proportion (analysis including 2-month window around index date)

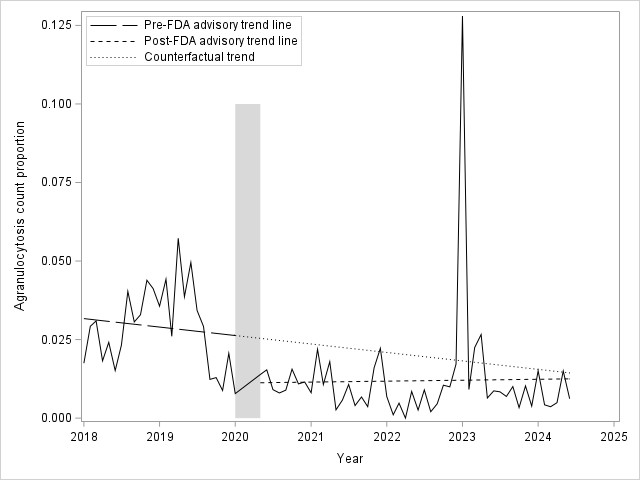

**β_1_:** -0.0002, **β_2_:** -0.015, **β_3_:** 0.0002, **F**: 4.71, **p-value:** 0.075

**Supplementary Figure 6:** Results of interrupted time series myocarditis count per month (analysis including 2-month window around index date)

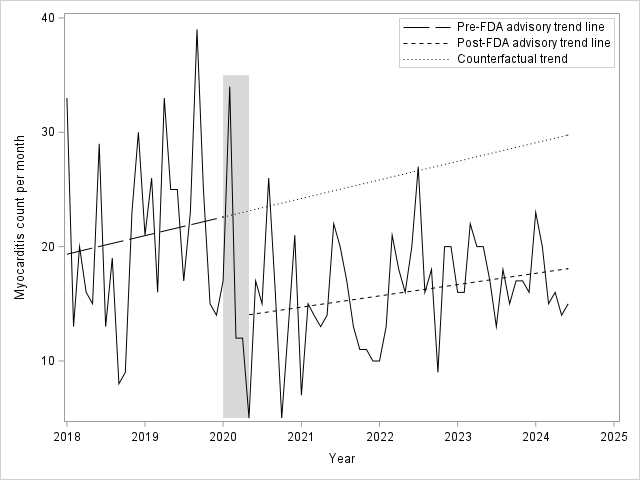

**β_1_:** 0.135, **β_2_:** -8.692, **β_3_:** -0.058, **F**: 1.81, **p-value:** 0.012

**Supplementary Figure 7:** Results of interrupted time series myocarditis count proportion (analysis including 2-month window around index date)

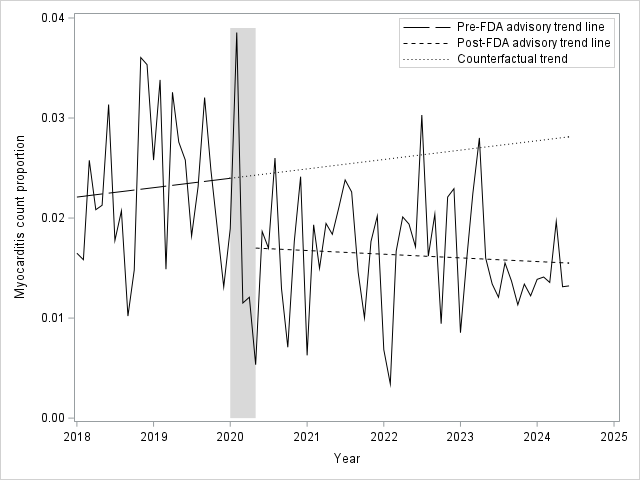

**β_1_:** 0.00007, **β_2_:** -0.007****, **β_3_**-0.0001, **F**: 2.20, **p-value:** 0.118

**Supplementary Figure 8:** Results of interrupted time series neutropenia count per month (analysis including 2-month window around index date)

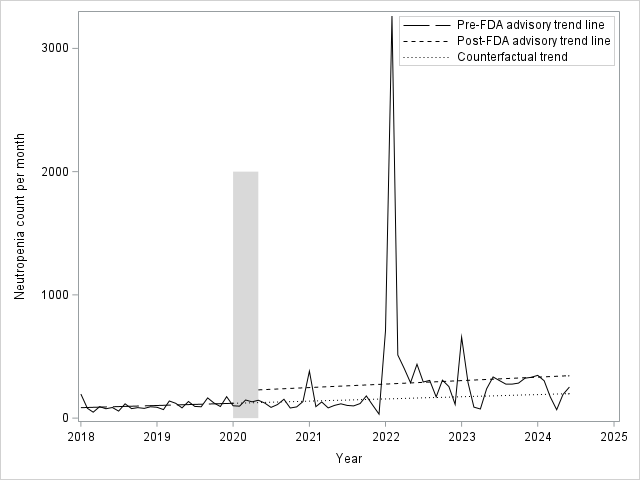

**β_1_:** 1.476, **β_2_:** 19.400, **β_3_:** 1.801, **F**: 0.14, **p-value:** 0.870

**Supplementary Figure 9:** Results of interrupted time series neutropenia count proportion (analysis including 2-month window around index date)

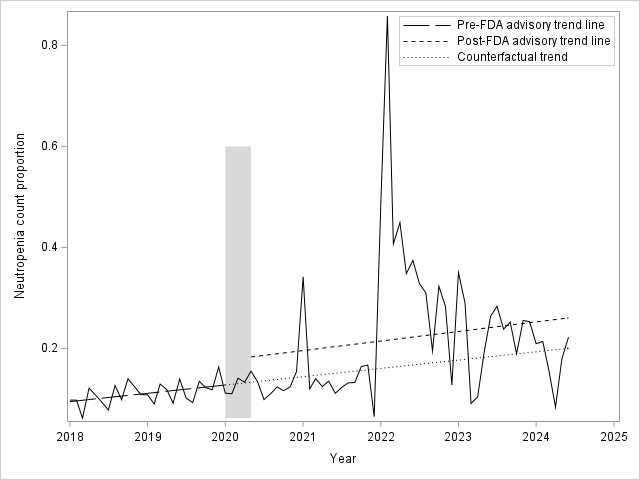

**β_1_:** 0.001, **β_2_:** 0.0354, **β_3_:** 0.0003, **F**: 0.39, **p-value:** 0.681
